# High levels of detection of non-pneumococcal species of *Streptococcus* in saliva from adults in the USA

**DOI:** 10.1101/2022.11.20.22282557

**Authors:** Maikel S. Hislop, Orchid M. Allicock, Darani A. Thammavongsa, Sidiya Mbodj, Allison Nelson, Albert C. Shaw, Daniel M. Weinberger, Anne L. Wyllie

**Author notes:** **Corresponding author: Anne Wyllie PhD**, Yale School of Public Health, LEPH 823, 60 College St, New Haven, CT 06510. Contributed equally (Maikel S. Hislop, Orchid M. Allicock). Co-senior authors (Daniel M. Weinberger, Anne L. Wyllie).

## Abstract

**Background:** While the sensitivity of detection of pneumococcal carriage can be improved by testing respiratory tract samples with qPCR, concerns have been raised regarding the specificity of this approach. We therefore investigated the reliability of the widely-used *lytA* qPCR assay when applied to saliva samples from older adults in relation to a more specific qPCR assay (*piaB)*.

**Methods:** During the autumn/winter seasons of 2018/2019 and 2019/2020, saliva was collected at multiple timepoints from 103 healthy adults aged 21-40 (n=34) and ≥64 (n=69) years. Following culture-enrichment, extracted DNA was tested using qPCR for *piaB* and *lytA*. By sequencing the variable region of *rpsB* (S2-typing), we identified the species of bacteria isolated from samples testing *lytA*-positive only.

**Results:** While 30/344 (8.7%) saliva samples (16.5% individuals) tested qPCR-positive for both *piaB* and *lytA*, 52 (15.1%) samples tested *lytA*-positive only. No samples tested *piaB*-positive only. Through extensive re-culture of the 32 *lytA*-positive samples collected in 2018/2019, we isolated 23 strains (from 8 samples, from 5 individuals) that were also qPCR-positive for only *lytA*. Sequencing determined that *Streptococcus mitis* and *Streptococcus infantis* were predominantly responsible for this *lytA*-positive qPCR signal.

**Conclusions:** We identified a comparatively large proportion of samples generating positive signals with the widely used *lytA*-qPCR and identified non-pneumococcal streptococcal species responsible for this signal. This highlights the importance of testing for the presence of multiple gene targets in tandem for reliable and specific detection of pneumococcus in respiratory tract samples.

## BACKGROUND

Upper respiratory tract carriage of *Streptococcus pneumoniae* is considered a prerequisite for invasive pneumococcal disease (IPD). Rates of carriage are highest in young children [1–4]. Carriage of pneumococcus is less commonly detected in older adults through a culture-based approach, despite a high incidence of disease in this age group [5–11]. Recent studies have demonstrated that when more sensitive methods are used, namely qPCR, higher rates of carriage in this at-risk age group can be detected [12,13], especially when using samples from oral sites [4,5,14–16].

For the detection of pneumococcus by qPCR, a suitable gene target must be selected to ensure both sensitive and specific identification; multiple gene targets further increases specificity, reducing false positive detection of pneumococcus [17]. However, the majority of molecular assays available for the detection of pneumococcus are developed using pure isolates, isolates in cell culture or from nasopharyngeal swabs. Thus, these assays do not account for the complicated microbial composition of the oropharynx/oral cavity and the potential for detection of related non-pneumococcal Streptococci [18,19].

The variability in sample types tested and detection methods applied (culture vs. molecular), leads to difficulty comparing results between settings. In the current study, we validated and evaluated previously reported methods [4,20] for qPCR testing of culture-enriched saliva. The frequent identification of non-pneumococcal *Streptococcus* strains in individuals testing positive for *lytA* alone highlights the importance of testing multiple gene targets (*lytA and piaB)* to obtain reliable detection.

## METHODS

### Study design and population

During the autumn/winter seasons (October to January) of 2018/2019 and 2019/2020, de-identified saliva samples were collected from healthy adults aged 21-40 years (community-dwelling) and ≥64 years (both community-dwelling and nursing home residents) as part of a study on influenza vaccination. Saliva was collected from study participants on the day of seasonal influenza vaccination (day 0) and on days 2, 7, 28 and 70. For the 2019/2020 study season, samples were not collected on day 2. Written informed consent was obtained from all participants, and the study was conducted in compliance with Good Clinical Practice and the Declaration of Helsinki of the World Medical Association. The study was approved by the Institutional Review Board of the Yale Human Research Protection Program (Protocol ID. 0409027018).

### Sample collection and processing

All saliva samples were self-collected by study participants under supervision by trained study personnel, placed on wet ice and transferred to the study laboratory at the Yale School of Public Health and processed within 4 hours of collection. On arrival at the lab, 100 μl of each sample was cultured on blood agar plates supplemented with gentamicin (10%) (Remel, Lenexa, KS) [5]. Following overnight incubation at 37ºC with 5% CO_2_, all bacterial growth was harvested from culture plates into brain heart infusion (BHI) supplemented with 10% glycerol and stored at -80°C until further analysis. These culture-plate harvests were considered to be culture-enriched for pneumococci.

### Detection of pneumococcus

Culture-enriched saliva samples were thawed on wet ice. DNA was extracted from 200 μl of each sample as previously described [21]. Each DNA template was tested in qPCR for pneumococcal genes *piaB* [3,16] and *lytA* [13]. The assays were carried out in 20 μl reaction volumes using SsoAdvanced Universal Probe Supermix (Biorad, USA), 2.5 μl of genomic DNA and primer/probe mixes (Iowa Black quenchers) for *piaB* (1 μl) and *lytA* (1.2 μl) at concentrations of 200 nM. DNA of *S. pneumoniae* serotype 19F strain #64 [22] was included as a positive control in every run. Assays were run on a CFX96 Touch (Biorad) under the following conditions: 95°C for 3 minutes, followed by 45 cycles of 98°C for 15 seconds and 60°C for 30 seconds. Samples were considered positive for pneumococcus when C_*T*_ values for both genes were ≤40 [23].

### Isolation of bacterial strains

Culture-enriched saliva samples qPCR-positive for only *lytA* were re-visited by culture in an attempt to isolate strains responsible for the *lytA* signal [16,23]. All strains isolated by re-culture were re-tested for *lytA* and *piaB* then stored at -80°C until further processing.

### S2-typing

The *lytA*-positive/*piaB*-negative strains were thawed on ice, cultured on blood agar plates using a 10 μl inoculating loop and incubated overnight at 37ºC with 5% CO_2_. Colonies were harvested using a 10 μl inoculating loop and DNA was extracted via the boilate method [21]. For each strain, the 408 bp variable region of the *rpsB* gene encoding the ribosomal protein S2 was amplified with conventional PCR using the primers previously described [24] with a modified protocol. Briefly, 2.5 μl of each DNA template was tested in 25 μl reaction volumes, consisting of 1 μl of each of the forward and reverse primers (10 μM each), 4 μl 5x Phusion HF Buffer, 0.2 μl Polymerase, 0.4 μl dNTP and 10.4 μl DNase free water. PCR conditions were as follows: 95°C for 15 minutes, then 40 cycles of 94°C for 30 seconds, 54°C for 1 minute and 72°C for 1 minute, followed by 60°C for 30 minutes. DNA of *S. pneumoniae* serotype 19F strain #64 was included as a positive control in every run. Since amplicons generated with the S2 primers can vary in size (species dependent) [24], amplicons between 400 and 450 bp were gel purified using the QIAquick Gel Extraction kit (QIAGEN). Each amplicon was adjusted to a final concentration of 60 ng, mixed with the S2F (4 μM) or S2R (4 μM) primer and sequenced by the Keck Biotechnology Resource Laboratory (Yale University, New Haven, CT, USA). Sequences generated were assembled using Geneious Prime 2022.1.1 (https://www.geneious.com) and cross-referenced with the reference dataset of 501 streptococcal S2-sequences for species annotation. Sequences were aligned using Geneious Prime 2022.1.1, employing the Clustal Omega algorithm [25].

### Statistical analysis

The study population was stratified into three age groups: younger adults (26–40 years old), older adults (60-79 years old) and the elderly (80-96). The risk factors for pneumococcal carrier status were investigated by generalized estimation equations, and the strength of association was expressed as odds ratios (ORs) with 95% confidence intervals (CI). Statistical analysis was performed using R version 3.6.1 with the following packages: dplyr [26], reshape2 [27], gee [28] and mgcv [29].

## RESULTS

### Population characteristics

A total of 344 samples was collected from 103 individuals over the course of the two study seasons. During the 2018/2019 study period, 197 samples were collected from 56 individuals. This includes 75 individuals aged 21-39 years enrolled from a workplace vaccination clinic and 122 individuals aged 64-95 years (51 enrolled from an aged-care living facility and 71 enrolled from a local health clinic). During the 2019/2020 study period, 147 samples were collected from 47 individuals, 42 from individuals aged 21-39 years enrolled from a workplace vaccination clinic and 105 from individuals aged 64-95 years (41 enrolled from an aged-care living facility and 64 enrolled from a local health clinic) (summarized in Table 1 and detailed per study period in Supplementary Table S1).

**Table 1.** Overall study participant demographics and PCR detection of *piaB* and *lytA* in saliva collected from (A) a workplace vaccination clinic, (B) an aged-care living facility, and (C) a local health clinic.

| Study year | Total |  |  |
| --- | --- | --- | --- |
| Study site | A | B | C |
| Total enrollment, n | 34 | 30 | 39 |
| Total number of samples collected, n | 117 | 92 | 135 |
| Average number of samples per person, (range) | 3 (1-5) | 3 (1-5) | 3 (1-5) |
| Age in years (median) | 21-39 (28) | 64-96 (88) | 65-88 (72) |
| Female | 23 | 16 | 21 |
| *piaB*+/*lytA*+ samples, n (%) | 14/117 (12.0%) | 2/92 (2.2%) | 13/135 (9.6%) |
| Period prevalence of pneumococcal carriage (*piaB*+ individuals), n (%) | 7/34 (20.6%) | 2/30 (6.7%) | 10/39 (25.6%) |
| *piaB*-/*lytA*+ samples, n (%) | 26/117 (22.2%) | 2/92 (2.2%) | 17/135 (12.6%) |

### Detection of Streptococcus pneumoniae

During the first study season (2018/2019) 16/197 (8.1%) culture-enriched saliva samples from 12/56 (21.4%) individuals tested qPCR-positive for both *piaB* and *lytA*, indicating the presence of pneumococcus. However, 27 (13.7%) samples from 14 (25.0%) individuals tested positive for *lytA* only. During the second study season (2019/2020) 14/147 (9.5%) culture-enriched saliva samples from 7/47 (14.9%) individuals tested qPCR positive for both *piaB* and *lytA*, and 18 (14.3%) samples from 9 (19.1%) individuals tested *lytA*-positive only. Several individuals were colonized with pneumococcus at multiple timepoints including one individual who was colonized throughout the sampling period (Figure 2). Pneumococcal colonization did not differ between the two study periods (OR 1.25, 95% confidence interval [CI] 0.42-3.74) nor was colonization dependent on sex (OR 1.24, CI 0.44-3.54). Individuals over 60 were less likely to be colonized with pneumococcus compared to younger study participants (63-80 year olds OR 0.71, CI 0.22-2.25; >80 year olds OR 0.27, CI 0.42-3.74).

**Figure 1.**
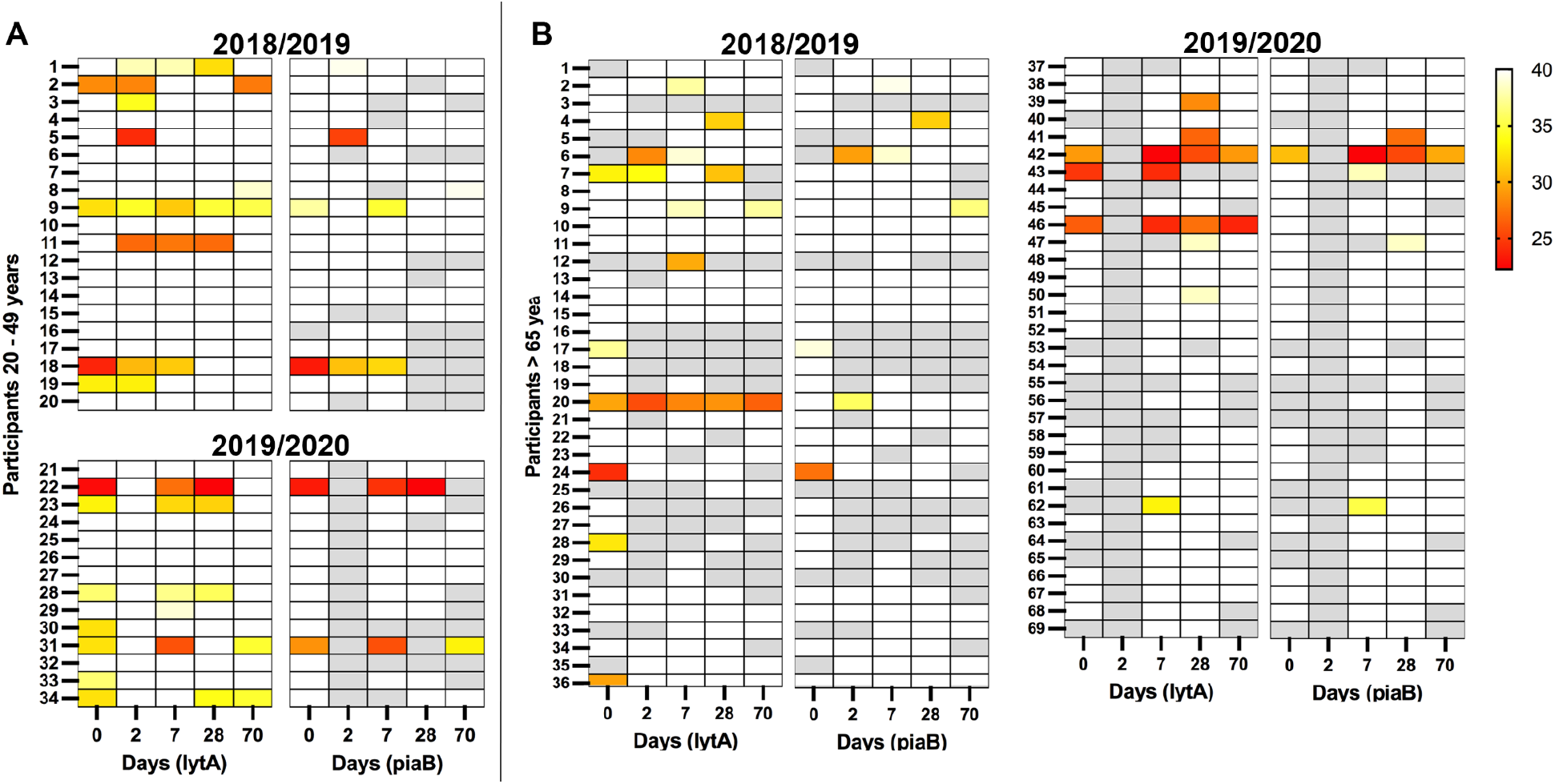
Detection of pneumococcal genes, *lytA* and *piaB* in saliva samples from healthy adults aged **A** 21-40 and **B** ≥65 years and older, collected during the autumn/winter seasons of 2018/2019 and 2019/2020. Each row represents a study participant, and each column is a sample. Darker colors indicate higher density of bacteria (lower PCR Ct value), white indicates no detection of the gene target, gray indicates the sample was not available for testing (either not collected or insufficient volume).

**Figure 2.**
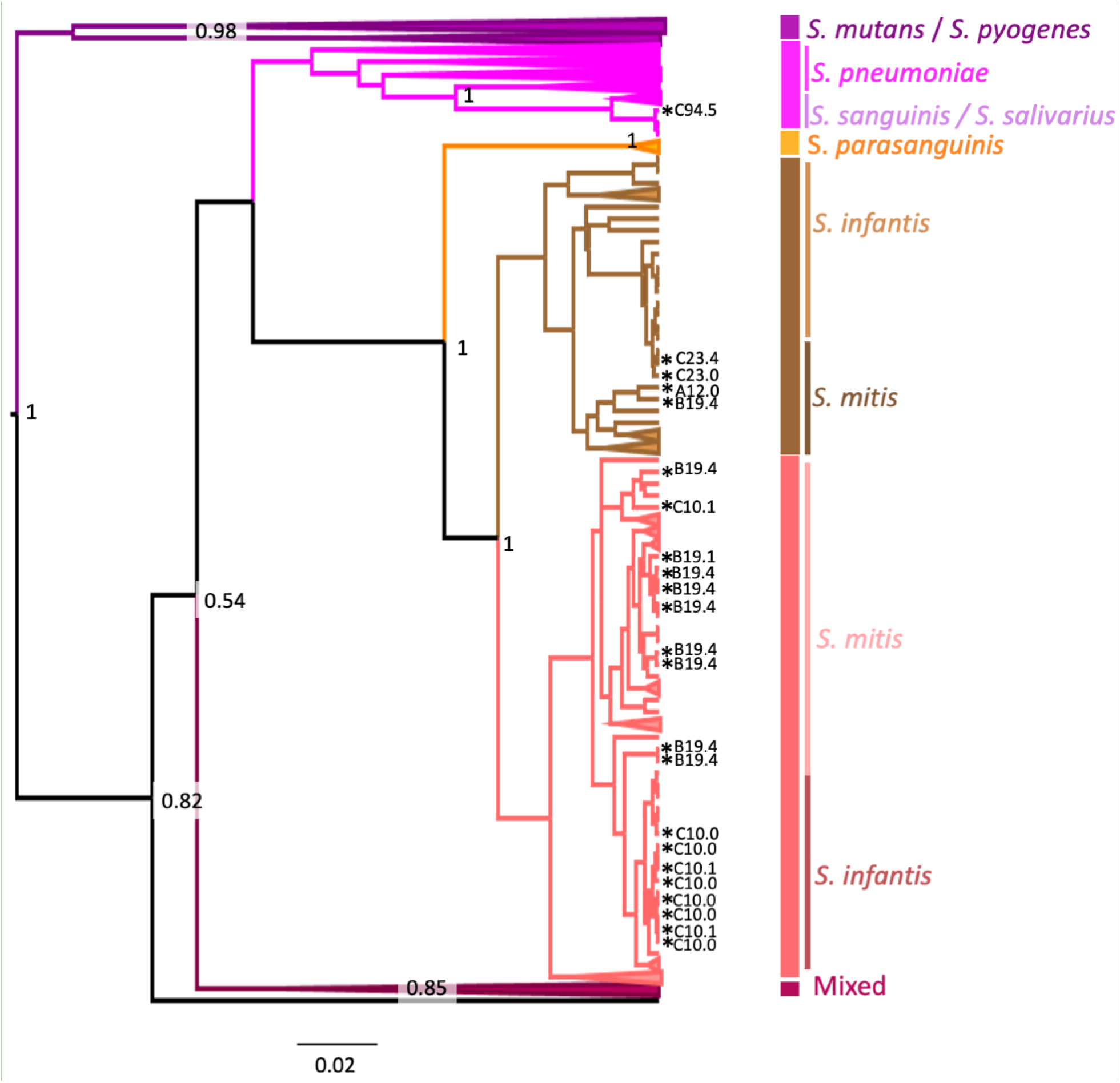
Bayesian Maximum Clade Credibility (MCC) tree inferred using rspB gene sequences (431 nt) derived from this study (n=23) along with sequences representing the main Streptococcus sp. (n=533). The mixed clade includes *S. gordonii, S. equi, S. anginosus, S. constellatus, S. canis* and *S. zooepidemicus*. Clade credibilities of 50% and over are indicated in black at the relevant nodes. Clades are coloured according to the main species found in the clade. Sequences from this study are marked with an asterisk (*). Phylogenetic analysis was performed using the GTR + G4 + I nucleotide model. Samples are labeled with their study ID consisting of study site (A = workplace vaccination clinic, B = aged-care living facility, C = local health clinic) and participant ID, followed by the time point at which the sample was collected (0 = day of influenza vaccination, 1 = 1 day after influenza vaccination, 4 = 28 days post-vaccination, 5 = 70 days post-vaccination).

### Identification of non-pneumococcal Streptococcus species

Through extensive re-culture of the *lytA*-positive samples from the first season, we isolated 23 non-pneumococcal *Streptococcus* strains (from 8 samples obtained from 5 individuals) which also tested qPCR-positive for only *lytA*. Colonization with *lytA-*positive non-pneumococcal *Streptococcus* strains did not differ between the two study periods, (OR 1.08, CI 0.39-2.99). Individuals over 60 were also less likely to be colonized with *lytA-*positive non-pneumococcal *Streptococcus* strains (63-80 year olds OR 0.38, CI 0.13-1.07; >80 year olds OR 0.23, CI 0.04-1.36).

Of the 23 isolates that were *lytA-*positive and *piaB-*negative, 11 (48%) isolates were identified by S2-typing as *S. mitis*, 11 (48%) isolates were identified as *S. infantis* and 1 (4%) was identified as *S. vestibularis* (see Supplementary Table S2). The *S. infantis* isolates were in the same clade or a sister branch of the reference taxon for *S. mitis, S. infantis* and *S. parasanguinis*.

## DISCUSSION

The current gold standard method for the detection of pneumococcal carriage is testing nasopharyngeal swabs by conventional culture. Updated World Health Organization recommendations made in 2013 by the Pneumococcal Carriage Working Group advised the inclusion of oropharyngeal swabs from adults when possible [30]. A number of studies have demonstrated that the sensitivity of carriage detection can be further improved when molecular methods are applied [15,31–34] to alternative sample types such as saliva [21,23]. With increased use of molecular methods however, there have been increased reports of confounding by non-pneumococcal species of *Streptococcus*, leading to uncertainty regarding the accuracy of culture-independent approaches [13,24,35]. While implementing previously established molecular methods [4] to standardize approaches for the detection of pneumococcus in saliva from healthy adults between geographic locales, we noted potential confounding with a large proportion of saliva samples, or 45 out of 103 individuals sampled, testing PCR positive for *lytA* but negative for *piaB*. This highlights the importance of testing for multiple specific gene targets when trying to accurately identify pneumococcal carriage from complex oral samples.

Targeting the *lytA* gene has become widely accepted for the identification of pneumococcus. However *lytA* homologues have been found in other streptococcal species [36], supporting the notion that detection of *lytA* alone can lead to pneumococcus misidentification [37]. The gene target *piaB* is specific for encapsulated pneumococci, but absent from other oral streptococci [38] and some unencapsulated pneumococci [24,39] meaning an underestimation of total pneumococcal carriage is possible. However, detection of both *lytA* and *piaB* genes by qPCR increases specificity and decreases false positive detection for *S. pneumoniae* [17]. Previous work has demonstrated concordance between *lytA* and *piaB* in the absence of unencapsulated or non-pneumococcal streptococci with *lytA* homologues [16]. Generally, when both *piaB* and *lytA* are detected in a sample at comparative Ct values, this is supportive of the presence of pneumococcus. However, it is not uncommon to see a stronger signal for *lytA*, indicating the co-presence of a non-pneumococcus species also carrying the *lytA* gene.

In line with this, we found that the majority of the confounding signal in the *lytA*-qPCR was caused by *Streptococcus mitis* and *Streptococcus infantis*. These results are in line with data from other studies detecting non-pneumococcal *Streptococcus* from oral samples [35,40], further supporting that the WHO recommendation of using non-culture-based molecular methods for pneumococcal detection should be revised to include multiple targets [35]. Studies reporting on carriage rates detected from oral sample types when using only one qPCR target must be cautious regarding the specificity and interpretation of their results. Primers and probes are typically developed on pure isolates or evaluated *in silico* on genomic sequences available in public databases. However, the majority of available sequences are from strains isolated from cases of disease. With commensals rarely implicated in disease, the extent of gene homologues in non-pneumococcus *Streptococcus* commensals is largely unknown. Thus, any assay developed for the detection of pneumococcal carriage, particularly in oral samples, should be thoroughly tested on both positive and negative samples to monitor potential of false-positivity. Accordingly, results from this study demonstrate the possibility of misidentification of streptococci when solely utilizing the *lytA* qPCR assay.

Understanding rates of carriage of pneumococcus in older adults is critical for evaluating vaccination strategies, both prior to and following their implementation. For this, nasopharyngeal swabbing has proved inefficient. Oral sample types improve carriage detection but need to be tested with care. Here, we detected a high frequency of samples testing qPCR-positive for *lytA* only, from which we isolated and identified non-pneumococcal species of *Streptococcus* responsible for this signal [24]. This underscores the importance of testing for the presence of multiple gene targets in tandem for reliable molecular detection of pneumococcus in respiratory tract samples; targeting only *lytA* may lead to an overestimation of true carriage rates.

## Data Availability

All data produced in the present study are available upon reasonable request to the authors

## DECLARATIONS

### Author’s contributions

ALW, ACS and DMW conceived the study. AN and ALW managed the study and collected the data. MSH, DAT and SM were responsible for and performed the assays. MSH, OMA, DMW and ALW performed the analyses and interpreted the data. MSH, OMA, and ALW drafted the manuscript. All authors amended and commented on the final manuscript.

## Acknowledgements

We gratefully acknowledge Denise Shepard, Gayle Mirto, Amy Shelton, Bridget Mignosa, David Nock, and Sandra Capelli for their efforts in supporting sample collection and the participants for their time and commitment to the study.

## SUPPLEMENTARY MATERIAL

Table S1. Study participant demographics and PCR detection of *piaB* and *lytA* genes in saliva collected from adults visiting (A) a workplace vaccination clinic, (B) an aged-care living facility, or (C) a local health clinic, by study period and overall.

Table S2. Identification of non-pneumococcal *Streptococcus* species generating positive signal in the *lytA*-PCR assay widely-used for pneumococcus detection, when testing saliva samples collected from adults visiting (A) a workplace vaccination clinic (21-40 year olds), (B) an aged-care living facility (≥64 year olds), and (C) a local health clinic (≥64 year olds).

## REFERENCES

1. Simell, B., Auranen, K., Käyhty, H., Goldblatt, D., Dagan, R., O’Brien, K.L., and Pneumococcal Carriage Group (2012). The fundamental link between pneumococcal carriage and disease. Expert Rev. Vaccines 11, 841–855.

2. Auranen, K., Mehtälä, J., Tanskanen, A., and S. Kaltoft M. (2009). Between-Strain Competition in Acquisition and Clearance of Pneumococcal Carriage—Epidemiologic Evidence From a Longitudinal Study of Day-Care Children. Am. J. Epidemiol. 171, 169– 176.

3. Wyllie, A.L., Wijmenga-Monsuur, A.J., van Houten, M.A., Bosch, A.A.T.M., Groot, J.A., van Engelsdorp Gastelaars, J., Bruin, J.P., Bogaert, D., Rots, N.Y., Sanders, E.A.M., et al. (2016). Molecular surveillance of nasopharyngeal carriage of Streptococcus pneumoniae in children vaccinated with conjugated polysaccharide pneumococcal vaccines. Sci. Rep. 6, 23809.

4. Wyllie, A.L., Rümke, L.W., Arp, K., Bosch, A.A.T., Bruin, J.P., Rots, N.Y., Wijmenga-Monsuur, A.J., Sanders, E.A.M., and Trzciński, K. (2016). Molecular surveillance on Streptococcus pneumoniae carriage in non-elderly adults; little evidence for pneumococcal circulation independent from the reservoir in children. Scientific Reports 6. 10.1038/srep34888.

5. Krone, C.L., Wyllie, A.L., van Beek, J., Rots, N.Y., Oja, A.E., Chu, M.L.J., Bruin, J.P., Bogaert, D., Sanders, E.A.M., and Trzciński, K. (2015). Carriage of Streptococcus pneumoniae in Aged Adults with Influenza-Like-Illness. PLOS ONE 10, e0119875. 10.1371/journal.pone.0119875.

6. Krone, C.L., van de Groep, K., Trzciński, K., Sanders, E.A.M., and Bogaert, D. (2014). Immunosenescence and pneumococcal disease: an imbalance in host–pathogen interactions. The Lancet Respiratory Medicine 2, 141–153.

7. Kolšek-šušteršič, M., Krasnič, A.B., Mioč, V., Paragi, M., and Rifel, J. (2017). Nasopharyngeal carriage of Streptococcus pneumoniae and serotypes indentified among nursing home residents in comparison to the elderly and patients younger than 65 years living in domestic environment. Slovenian Journal of Public Health 56, 172–178. 10.1515/sjph-2017-0023.

8. Kwetkat, A., Pfister, W., Pansow, D., Pletz, M.W., Sieber, C.C., and Hoyer, H. (2018). Naso- and oropharyngeal bacterial carriage in nursing home residents: Impact of multimorbidity and functional impairment. PLoS One 13, e0190716.

9. Milucky, J., Carvalho, M. de G., Rouphael, N., Bennett, N.M., Talbot, H.K., Harrison, L.H., Farley, M.M., Walston, J., Pimenta, F., Lessa, F.C., et al. (2019). Streptococcus pneumoniae colonization after introduction of 13-valent pneumococcal conjugate vaccine for US adults 65 years of age and older, 2015–2016. Vaccine 37, 1094–1100.

10. van Beek, J., Veenhoven, R.H., Bruin, J.P., van Boxtel, R.A.J., de Lange, M.M.A., Meijer, A., Sanders, E.A.M., Rots, N.Y., and Luytjes, W. (2017). Influenza-like Illness Incidence Is Not Reduced by Influenza Vaccination in a Cohort of Older Adults, Despite Effectively Reducing Laboratory-Confirmed Influenza Virus Infections. The Journal of Infectious Diseases 216, 415–424. 10.1093/infdis/jix268.

11. Satzke, C., Dunne, E.M., Porter, B.D., Klugman, K.P., Kim Mulholland, E., and PneuCarriage project group (2015). The PneuCarriage Project: A Multi-Centre Comparative Study to Identify the Best Serotyping Methods for Examining Pneumococcal Carriage in Vaccine Evaluation Studies. PLOS Medicine 12, e1001903. 10.1371/journal.pmed.1001903.

12. Turner, P., Hinds, J., Turner, C., Jankhot, A., Gould, K., Bentley, S.D., Nosten, F., and Goldblatt, D. (2011). Improved detection of nasopharyngeal cocolonization by multiple pneumococcal serotypes by use of latex agglutination or molecular serotyping by microarray. J. Clin. Microbiol. 49, 1784–1789.

13. Carvalho, M. da G.S., Tondella, M.L., McCaustland, K., Weidlich, L., McGee, L., Mayer, L.W., Steigerwalt, A., Whaley, M., Facklam, R.R., Fields, B., et al. (2007). Evaluation and improvement of real-time PCR assays targeting lytA, ply, and psaA genes for detection of pneumococcal DNA. J. Clin. Microbiol. 45, 2460–2466.

14. Tramuto, F., Amodio, E., Calamusa, G., Restivo, V., Costantino, C., Vitale, F., and The BINOCOLO Group (2017). Pneumococcal Colonization in the Familial Context and Implications for Anti-Pneumococcal Immunization in Adults: Results from the BINOCOLO Project in Sicily. Int. J. Mol. Sci. 18. 10.3390/ijms18010105.

15. Branche, A.R., Yang, H., Java, J., Holden-Wiltse, J., Topham, D.J., Peasley, M., Frost, M.R., Nahm, M.H., and Falsey, A.R. (2018). Effect of prior vaccination on carriage rates of Streptococcus pneumoniae in older adults: A longitudinal surveillance study. Vaccine 36, 4304–4310.

16. Trzciński, K., Bogaert, D., Wyllie, A., Chu, M.L.J., van der Ende, A., Bruin, J.P., van den Dobbelsteen, G., Veenhoven, R.H., and Sanders, E.A.M. (2013). Superiority of Trans-Oral over Trans-Nasal Sampling in Detecting Streptococcus pneumoniae Colonization in Adults. PLoS ONE 8, e60520. 10.1371/journal.pone.0060520.

17. Varghese, R., Jayaraman, R., and Veeraraghavan, B. (2017). Current challenges in the accurate identification of Streptococcus pneumoniae and its serogroups/serotypes in the vaccine era. J. Microbiol. Methods 141, 48–54.

18. Stearns, J.C., Davidson, C.J., McKeon, S., Whelan, F.J., Fontes, M.E., Schryvers, A.B., Bowdish, D.M.E., Kellner, J.D., and Surette, M.G. (2015). Culture and molecular-based profiles show shifts in bacterial communities of the upper respiratory tract that occur with age. ISME J. 9, 1268.

19. Ganaie, F., Branche, A.R., Peasley, M., Rosch, J.W., and Nahm, M.H. (2022). Effect of Oral Streptococci Expressing Pneumococcus-like Cross-Reactive Capsule Types on World Health Organization Recommended Pneumococcal Carriage Detection Procedure. Clin. Infect. Dis. 75, 647–656.

20. Krone, C.L., van Beek, J., Wyllie, A., Rots, N., Oja, A., Chu, M.L., Akenda, M.L., Bogaert, D., Sanders, E.A.M., and Trzcinski, K. (2013). High rates of Streptococcus pneumoniae carriage in saliva of elderly. Host and microbe characteristics of pneumococcal colonization in elderly, 87.

21. Wyllie, A.L., Mbodj, S., Thammavongsa, D.A., Hislop, M.S., Yolda-Carr, D., Waghela, P., Nakahata, M., Watkins, A.E., Vega, N.J., York, A., et al. (2022). Persistence of pneumococcal carriage among older adults in the community despite COVID-19 mitigation measures. bioRxiv. 10.1101/2022.06.28.22276654.

22. Tóthpál, A., Desobry, K., Joshi, S.S., Wyllie, A.L., and Weinberger, D.M. (2019). Variation of growth characteristics of pneumococcus with environmental conditions. BMC Microbiol. 19, 304.

23. Wyllie, A.L., Chu, M.L.J.N., Schellens, M.H.B., van Engelsdorp Gastelaars, J., Jansen, M.D., van der Ende, A., Bogaert, D., Sanders, E.A.M., and Trzciński, K. (2014). Streptococcus pneumoniae in saliva of Dutch primary school children. PLoS One 9, e102045.

24. Wyllie, A.L., Pannekoek, Y., Bovenkerk, S., van Engelsdorp Gastelaars, J., Ferwerda, B., van de Beek, D., Sanders, E.A.M., Trzciński, K., and van der Ende, A. (2017). Sequencing of the variable region of rpsB to discriminate between Streptococcus pneumoniae and other streptococcal species. Open Biol. 7. 10.1098/rsob.170074.

25. Sievers, F., Wilm, A., Dineen, D., Gibson, T.J., Karplus, K., Li, W., Lopez, R., McWilliam, H., and Remmert, M. ding, JSO, Thompson, JD, and Higgins, DG (2011) Fast, scalable generation of high-quality protein multiple sequence alignments using Clustal Omega. Mol. Syst. Biol. 7, 1–6.

26. Wickham, H., François, R., Henry, L., and Müller, K. dplyr: A grammar of data manipulation. R package version 0.4.

27. Wickham, H. (2007). Reshaping Data with the reshape Package. J. Stat. Softw. 21, 1–20.

28. Carey, V.J., Lumley, T., and Ripley, B. Gee: generalized estimation equation solver. 2012.. R-project. org/package= gee. R package version.

29. Wood, S.N. (2017). Generalized Additive Models: An Introduction with R (CRC Press/Taylor & Francis Group).

30. Satzke, C., Turner, P., Virolainen-Julkunen, A., Adrian, P.V., Antonio, M., Hare, K.M., Henao-Restrepo, A.M., Leach, A.J., Klugman, K.P., Porter, B.D., et al. (2013). Standard method for detecting upper respiratory carriage of Streptococcus pneumoniae: Updated recommendations from the World Health Organization Pneumococcal Carriage Working Group. Vaccine 32, 165–179. 10.1016/j.vaccine.2013.08.062.

31. Miellet, W.R., van Veldhuizen, J., Litt, D., Mariman, R., Wijmenga-Monsuur, A.J., Badoux, P., Nieuwenhuijsen, T., Thombre, R., Mayet, S., Eletu, S., et al. (2022). It Takes Two to Tango: Combining Conventional Culture With Molecular Diagnostics Enhances Accuracy of Streptococcus pneumoniae Detection and Pneumococcal Serogroup/Serotype Determination in Carriage. Frontiers in Microbiology 13. 10.3389/fmicb.2022.859736.

32. Ricketson, L.J., Lidder, R., Thorington, R., Martin, I., Vanderkooi, O.G., Sadarangani, M., and Kellner, J.D. (2021). PCR and Culture Analysis of Streptococcus pneumoniae Nasopharyngeal Carriage in Healthy Children. Microorganisms 9. 10.3390/microorganisms9102116.

33. Carvalho, M.D.G., Pimenta, F.C., Jackson, D., Roundtree, A., Ahmad, Y., Millar, E.V., O’Brien, K.L., Whitney, C.G., Cohen, A.L., and Beall, B.W. (2010). Revisiting pneumococcal carriage by use of broth enrichment and PCR techniques for enhanced detection of carriage and serotypes. J. Clin. Microbiol. 48, 1611–1618.

34. Saukkoriipi, A., Leskelä, K., Herva, E., and Leinonen, M. (2004). Streptococcus pneumoniae in nasopharyngeal secretions of healthy children: comparison of real-time PCR and culture from STGG-transport medium. Mol. Cell. Probes 18, 147–153.

35. Ganaie, F., Branche, A.R., Peasley, M., Rosch, J.W., and Nahm, M.H. (2021). Oral streptococci expressing pneumococci-like cross-reactive capsule types can affect WHO recommended pneumococcal carriage procedure. Clin. Infect. Dis. 10.1093/cid/ciab1003.

36. Whatmore, A.M., Efstratiou, A., Paul Pickerill, A., Broughton, K., Woodard, G., Sturgeon, D., George, R., and Dowson, C.G. (2000). Genetic Relationships between Clinical Isolates of Streptococcus pneumoniae, Streptococcus oralis, and Streptococcus mitis : Characterization of “Atypical” Pneumococci and Organisms Allied to S. mitis Harboring S. pneumoniae Virulence Factor-Encoding Genes. Infection and Immunity 68, 1374–1382. 10.1128/iai.68.3.1374-1382.2000.

37. Simões, A.S., Tavares, D.A., Rolo, D., Ardanuy, C., Goossens, H., Henriques-Normark, B., Linares, J., de Lencastre, H., and Sá-Leão, R. (2016). lytA-based identification methods can misidentify Streptococcus pneumoniae. Diagn. Microbiol. Infect. Dis. 85, 141–148.

38. Whalan, R.H., Funnell, S.G.P., Bowler, L.D., Hudson, M.J., Robinson, A., and Dowson, C.G. (2006). Distribution and genetic diversity of the ABC transporter lipoproteins PiuA and PiaA within Streptococcus pneumoniae and related streptococci. J. Bacteriol. 188, 1031– 1038.

39. Tavares, D.A., Simões, A.S., Bootsma, H.J., Hermans, P.W., de Lencastre, H., and Sá-Leão, R. (2014). Non-typeable pneumococci circulating in Portugal are of cps type NCC2 and have genomic features typical of encapsulated isolates. BMC Genomics 15, 863.

40. Sadowy, E., and Hryniewicz, W. (2020). Identification of Streptococcus pneumoniae and other Mitis streptococci: importance of molecular methods. Eur. J. Clin. Microbiol. Infect. Dis. 39, 2247–2256.

