## Supplementary for "High levels of detection of non-pneumococcal species of *Streptococcus* in saliva from adults in the USA"

[#]Contributed equally
*Co-senior authors

**Table S1.** Study participant demographics and PCR detection of *piaB* and *lytA* genes in saliva collected from adults visiting (A) a workplace vaccination clinic, (B) an aged-care living facility, or (C) a local health clinic, by study period and overall.

| **Study year** | **2018/2019** | | | **2019/2020** | | | **Total** | | |
|---|---|---|---|---|---|---|---|---|---|
| Study site | A | B | C | A | B | C | A | B | C |
| Total enrollment, n | 20 | 16 | 20 | 14 | 14 | 19 | 34 | 30 | 39 |
| Total number of samples collected, n | 75 | 51 | 71 | 42 | 41 | 64 | 117 | 92 | 135 |
| Average number of samples per person, (range) | 4 (1-5) | 3 (1-5) | 4 (1-5) | 3 (1-4) | 3 (1-4) | 3 (1-4) | 3 (1-5) | 3 (1-5) | 3 (1-5) |
| Age in years (median) | 21-39 (29) | 64-95 (89) | 65-88 (73) | 23-35 (27) | 66-96 (82) | 65-85 (71) | 21-39 (28) | 64-96 (88) | 65-88 (72) |
| Female | 13 | 8 | 11 | 10 | 8 | 10 | 23 | 16 | 21 |
| *piaB+/lytA+ samples, n (%)* | 8/75 (10.7%) | 1/51 (2.0%) | 7/71 (9.9%) | 6/42 (14.3%) | 1/64 (1.6%) | 7/64 (10.9%) | 14/117 (12.0%) | 2/92 (2.2%) | 13/135 (9.6%) |
| Period prevalence of pneumococcal carriage (*piaB*+ individuals), n (%) | 5/20 (25.0%) | 1/16 (6.3%) | 6/20 (30.0%) | 2/14 (14.3%) | 1/14 (7.1%) | 4/19 (21.0%) | 7/34 (20.6%) | 2/30 (6.7%) | 10/39 (25.6%) |
| *piaB-/lytA+ samples, n (%)* | 14/75 (18.7%) | 2/51 (3.9%) | 11/71 (15.5%) | 12/42 (28.6%) | 0/64 | 6/64 (9.4%) | 26/117 (22.2%) | 2/92 (2.2%) | 17/135 (12.6%) |

**Table S2**. Identification of non-pneumococcal *Streptococcus* species generating positive signal in the *lytA*-PCR assay widely-used for pneumococcus detection, when testing saliva samples collected from adults visiting (A) a workplace vaccination clinic (21-40 year olds), (B) an aged-care living facility (≥64 year olds), and (C) a local health clinic (≥64 year olds).

| Setting | Study ID | Timepoint (day) | Isolate ID | Species identified by S2-typing (tree alignment) | NCBI blast of *rpsB* sequence generated by S2-typing |
|---|---|---|---|---|---|
| A | 12 | 0 | 5 | *S. infantis* | *S. oralis/oral/VT* |
| B | 19 | 2 | 2 | *S. mitis* | *S.mitis* |
| B | 19 | 70 | 4 | *S. mitis* | *S. mitis* |
| B | 19 | 70 | 5 | *S. mitis* | *S. mitis/S. pseudopneumoniae* |
| B | 19 | 70 | 6 | *S. mitis* | *S. mitis* |
| B | 19 | 70 | 8 | *S. mitis* | *S. mitis/S. pseudopneumoniae* |
| B | 19 | 70 | 1.1 | *S. mitis* | *S. mitis/S. oralis* |
| B | 19 | 70 | 4.1 | *S. mitis* | *S. mitis/S. sp oral taxon* |
| B | 19 | 70 | 5.1 | *S. mitis* | *S. mitis/S. pseudopneumoniae* |
| B | 19 | 70 | 7.1 | *S. mitis* | *S. mitis/S. pseudopneumoniae* |
| B | 19 | 70 | 8.1 | *S. mitis* | *S. mitis/S. pseudopneumoniae* |
| C | 10 | 0 | 2 | *S. infantis* | *S. sp oral taxon/S. mitis* |
| C | 10 | 0 | 3 | *S. infantis* | *S. sp oral taxon/S. mitis* |
| C | 10 | 0 | 8 | *S. infantis* | *S. sp oral taxon/S. mitis* |
| C | 10 | 0 | 2.1 | *S. infantis* | *S. sp. oral taxon/S. mitis* |
| C | 10 | 0 | 3.1 | *S. infantis* | *S. sp. oral taxon/S. mitis* |
| C | 10 | 0 | 8.1 | *S. infantis* | *S. sp. oral taxon/S. mitis* |
| C | 10 | 2 | 4 | *S. mitis* | *S. mitis/S. pseudopneumoniae* |
| C | 10 | 2 | 4.1 | *S. infantis* | *S. sp. oral taxon/S. mitis* |
| C | 10 | 2 | 5.1 | *S. infantis* | *S. mitis/S. pseudopneumoniae* |
| C | 23 | 0 | 1.2 | *S. infantis* | *S. oralis/S. sp. oral taxon* |
| C | 23 | 70 | 7.1 | *S. infantis* | *S. oralis/S. sp. oral taxon* |
| C | 94 | 0 | 5 | *S. sanguinis* | *S. vestibularis/S. thermophilus/S. salivarius* |
